# Multimodal Large Language Models are Generalist Medical Image Interpreters

**DOI:** 10.1101/2023.12.21.23300146

**Authors:** Tianyu Han, Lisa C. Adams, Sven Nebelung, Jakob Nikolas Kather, Keno K. Bressem, Daniel Truhn

## Abstract

Medicine is undergoing a transformation with the integration of Artificial Intelligence (AI). Traditional AI models, though clinically useful and often matching or surpassing expert clinicians in specific tasks, face a scalability challenge due to the necessity of developing individual models for each task. Therefore, there is a push towards foundation models that are applicable to a wider set of tasks. Our study showcases how non-domain-specific, publicly available vision-language models can be employed as general foundation models for medical applications. We test our paradigm across four medical disciplines - pathology, dermatology, ophthalmology, and radiology - focusing on two use-cases within each discipline. We find that our approach beats existing pre-training methods and is competitive to domain-specific foundation models that require vast amounts of domain-specific training images. We also find that large vision-language models are data efficient and do not require large annotated datasets to reach competitive performance. This allows for the development of new or improved AI models in areas of medicine where data is scarce and will accelerate medical progress towards true multimodal foundation models.

## Introduction

Medicine is in the process of being transformed by Artificial Intelligence (AI). There is now ample evidence that specialized AI models have clinical use and can reach - or even surpass - the performance of expert clinicians in narrow tasks^1–7^. However, there is an inherent limitation for the development and application of such models: they need to be trained for each task separately. With thousands of potential applications, this is unsustainable, limiting the scalability and practicality of such specialized models.

This is why there is a shift in focus towards foundation models which are envisioned to be applicable to a wide range of tasks^8,9^. Recently, first steps towards this goal have been taken: Zhou et al. demonstrated the performance of a foundation model for generalizable disease detection from retinal images^10^. This model has been trained on over a million retinal images and can be applied to both fundoscopies and optical coherence tomography of the retina to diagnose a range of ocular diseases. Similarly, Huang et al. trained a foundation model for pathology image analysis by making use of the histopathological images that are available on X (formerly Twitter) together with accompanying texts^11,12^. Both of these models have two things in common: they are each applicable to a wider, but still limited domain and they require a large dataset of training images from that particular domain. In that sense, these models are more foundational than the specialized AI models, but they still fall short of being true foundational models^9,13^.

We take the next step by demonstrating that it is not necessary to train such domain-specific foundation models for downstream tasks. We instead employ a large publicly available vision-language model that has been trained on non-domain-specific data from the internet as a general foundation model. Large vision-language models can answer complex medical questions almost on par with human experts^14,15^ (**Figure 1**). We test this model in four medical fields heavily reliant on image classification: pathology, dermatology, ophthalmology, and radiology. In particular, we select two use-cases for each of these disciplines and examine how the large vision-language model internally represents these images and whether this internal representation allows distinguishing between various medical subclasses for downstream classification.

**Figure 1:**
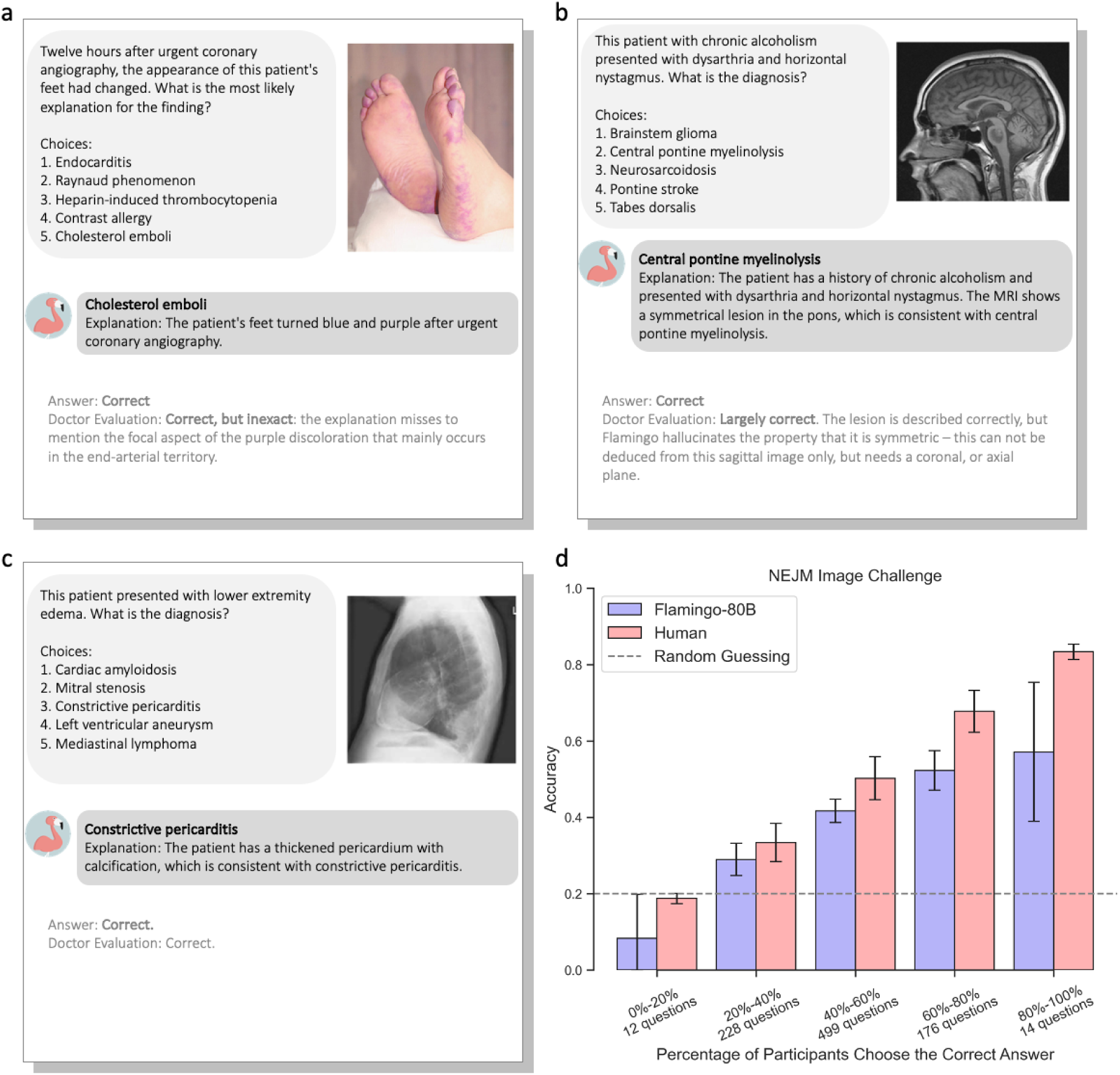
Performance of the multimodal LLM with 80 billion parameters on the NEJM Image Challenge Cases. (a)-(c): Selected NEJM cases correctly answered by the multimodal 80B LLM. The model provided the answer along with an explanation that was checked by a board-certified radiologist with 12 years of experience. (d): performance of Flamingo-80B in the NEJM challenge as compared to non-selective human participants. Bars indicate accuracy means; vertical lines indicate standard deviations.NEJM - The New England Journal of Medicine.

By demonstrating the competitive performance of this large vision-language model, we solve a serious problem that has so far impeded medical research: there is no need for large annotated datasets anymore if you aim to train a medical foundation model. Instead, you can utilize existing vision-language models and fine-tune them to your downstream task with limited labeled data.

We thus deliver proof that the era of general foundation models in medicine has already begun and that these models can accelerate medical progress by democratizing access to foundation models.

## Methods

### Ethics Approval

This study was conducted in accordance with the tenets of the Declaration of Helsinki and was approved by the local institutional review board (EK259/22).

### Patient Cohorts and Imaging Data

In this study, we systematically examined medical imaging datasets across four key medical disciplines: pathology, dermatology, ophthalmology, and radiology. We conducted two specific image classification tasks within each discipline, resulting in a total of eight distinct tasks (**T**), see **Figure 2a** and **Table 1**:

**Figure 2:**
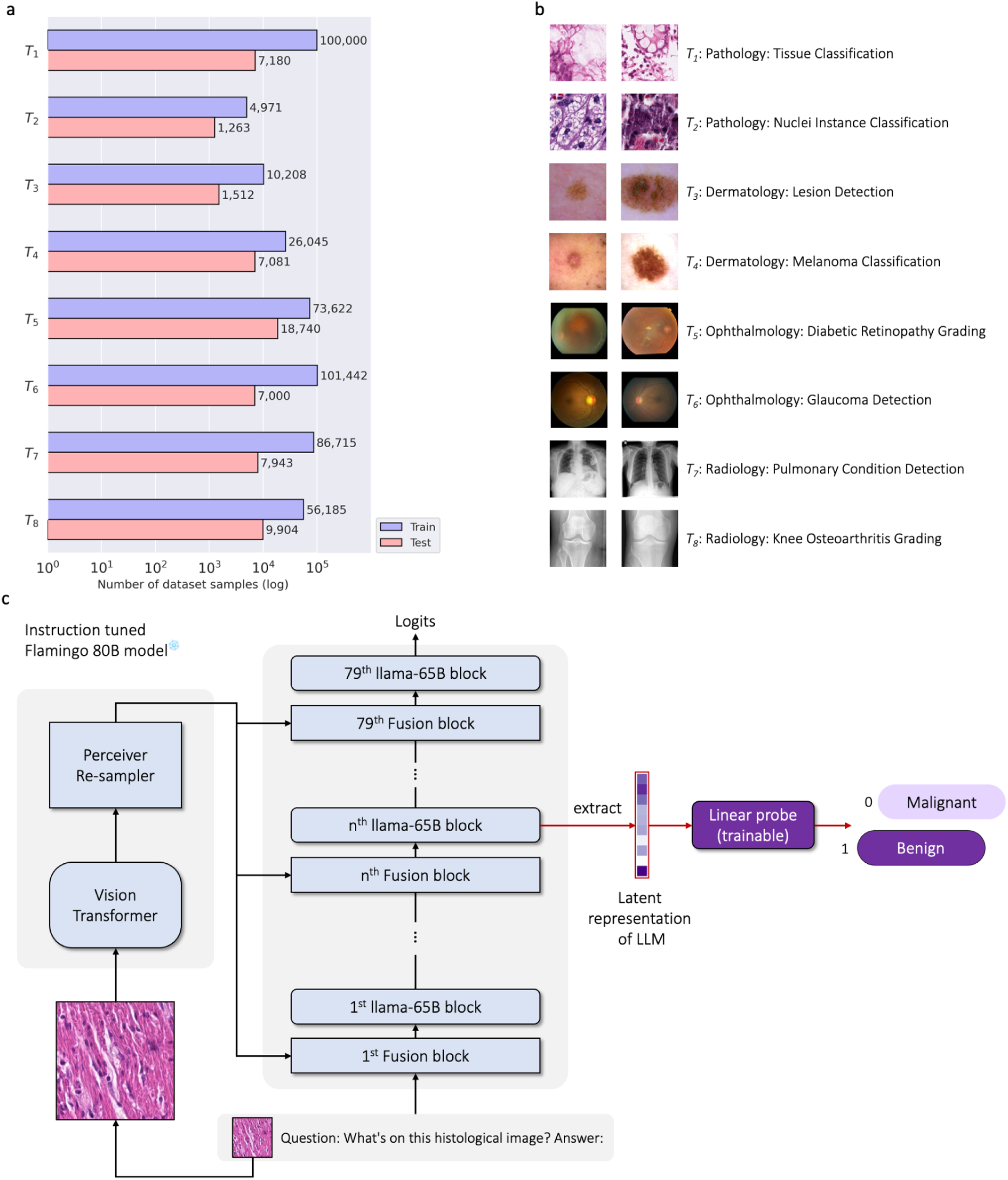
Setup of Experiments. (a)-(b): Flamingo (80B and 9B) models were evaluated on eight image classification tasks of four medical imaging domains. (c): Visualization of the probing of internal states used for the classification. Both vision and LLM-trained weights are frozen during probing (colored in light blue).

**Table 1:** Details on eight image classification tasks.

| Task | Type | Datasets | Training Set |  | Testing Set |  |
| --- | --- | --- | --- | --- | --- | --- |
|  |  |  | Images | Patients | Images | Patients |
| T1 | Histopathology | NCT-CRC-HE-100K<sup>16</sup><br>CRC-VAL-HE-7K<sup>16</sup> | 100,000 | 136 | 7,180 | 50 |
| T2 | Histopathology | PanNuke<sup>17</sup> | 4,971 | N/A | 1,263 | N/A |
| T3 | Dermatology | HAM10000<sup>19</sup><br>ISIC 2018<sup>18</sup> | 10,208 | N/A | 1,512 | N/A |
| T4 | Dermatology | SIIM-ISIC 2020<sup>20</sup> | 26,045 | 1,644 | 7,081 | 412 |
| T5 | Ophthalmology | EyePACS<sup>21</sup> | 73,622 | 36,811 | 15,078 | 7,539 |
|  |  | APTOS-2019<sup>22</sup> | 0 | 0 | 3,662 | N/A |
| T6 | Ophthalmology | AIROGS<sup>23</sup> | 101,442 | 54,274 | 0 | 0 |
|  |  | ODIR-2019<sup>24</sup> | 0 | 0 | 7,000 | 3,500 |
| T7 | Radiology | PadChest<sup>26</sup> | 86,715 | 59,975 | 7,943 | 7,272 |
| T8 | Radiology | OAI<sup>27,28</sup> | 39,921 | 3,831 | 7,108 | 677 |
|  |  | MOST<sup>27,28</sup> | 16,264 | 2,594 | 2,796 | 418 |

#### Tissue Classification in Histopathology Images (T1)

Using the NCT-CRC-HE-100K dataset, this task includes histological imaging data from 136 colorectal cancer patients. Following the dataset partitioning proposed by Kather et al^16^, we formed a training set of 100,000 image patches from 86 patients and a test set of 7,180 patches from 50 patients. Each patch, measuring 224×224 pixels, is classified into one of nine tissue categories: adipose tissue, background, debris, lymphocytes, mucus, smooth muscle, normal colonic mucosa, cancer-associated stroma, and colorectal adenocarcinoma epithelium^16^.

#### Nuclear Classification in Histopathology Images (T2)

This task uses the PanNuke dataset, which contains 7,558 pan-cancer images from 19 different organ types^17^. These images, which were annotated by Gamper et al., include various nuclear categories such as neoplastic, inflammatory, connective, epithelial, and dead tissue, including both apoptotic and necrotic cells.

#### Lesion Detection in Dermatology (T3)

For this task, we utilized the 2018 International Skin Imaging Collaboration (ISIC) Challenge dataset, comprising 10,208 training and 1,512 testing images of various skin lesions. Classifications include melanoma, basal cell carcinoma, and several other lesion types, as detailed in the work by Tschandl et al^18,19^.

#### Melanoma Classification in Dermatology (T4)

Derived from the ISIC 2020 Challenge, this task includes dermatology data with images labeled as benign or malignant^20^. The dataset, which differs from the 2018 challenge, includes 26,045 images for training and 7,081 for testing, stratified by patient (1,644 patients for training, 412 for testing).

#### Diabetic Retinopathy Grading in Fundoscopic Images (T5)

We sourced data from the 2015 EyePACS Diabetic Retinopathy Detection Challenge^21^ and the APTOS-2019 Blindness Detection Challenge^22^, totaling 88,700 fundoscopies from 44,350 patients. The combined dataset was divided into 73,622 training images (only EyePACS) and 18,740 testing images (from EyePACS (7,539 patients) and APTOS-2019).

#### Glaucoma Detection in Fundoscopic Images (T6)

This task incorporates data from the AIROGS^23^ and ODIR-2019^24^ challenges, resulting in a large dataset of 101,442 fundoscopies from 54,274 patients for training and 7,000 fundoscopies from 3,500 patients for testing.

#### Lung Disease Detection in Chest Radiographs in Radiology (T7)

Using the ‘PadChest’ cohort, this task focuses on radiology data with 86,715 chest radiographs from 59,975 patients for training and 7,943 radiographs from 7,272 patients for testing^25,26^. The dataset includes 174 radiographic findings and 19 radiological diagnoses^26^.

#### Osteoarthritis Grading in Knee Radiographs in Radiology (T8)

Employing data from the Osteoarthritis Initiative (OAI) and the Multicenter Osteoarthritis Study (MOST), this task involves grading osteoarthritis in knee radiographs^27,28^. Following the methodology of Han et al.^4^, we constructed a dataset with 56,185 training images from 6,425 patients and 9,904 testing images from 1,095 patients.

### NEJM Image Challenge Benchmarking

In this study, we collected 931 clinical cases from the New England Journal of Medicine (NEJM) Image Challenge from October 2005 to August 2023. Each case presented a medical image accompanied by a short text describing the clinical context, culminating in a specific question such as “What is the diagnosis?” (see **Figure S1** for an example). We provided five possible answers for each case and tasked DeepMind’s Flamingo model with selecting the correct answer.^29^ The dataset covered a wide range of medical fields, including pathology, dermatology, ophthalmology, and radiology, providing a comprehensive mix of medical imaging data. Statistics on the number of correct answers provided by NEJM readers were used to stratify the difficulty of the questions into five equal intervals according to the percentage of correct answers provided by human readers.^3^

We used a few-shot, in-context learning approach to test Flamingo on the NEJM cases.^30^ This involved providing the first two cases from the dataset (dated October 13th and 20th, 2005) to the model (**Figure S2**). The remaining 929 cases were then used as a test set to assess the model’s ability to interpret medical images across different disciplines.

### Vision-Language Model

We used the open-source Flamingo architecture, ^31^ which was trained by Hugging Face M4 and is available in two sizes: Flamingo-80B with 80 billion parameters and Flamingo-9B with 9 billion parameters. Both models are vision-language models that accept text interleaved with images and output free-form text. Flamingo combines a pre-trained large language model (LLM, LLaMA-65B for Flamingo-80B and Llama-7B for Flamingo-9B^32^) and a pre-trained Vision Transformer (ViT, 632M parameters^33^) via a transformer-based mapper (Perceiver Sampler^34^). To fuse vision and text signals, Flamingo uses cross-attention layers interleaved with LLM residual blocks (see **Figure 2c**). LLaMA-65B was pre-trained on 1.4 trillion tokens from publicly available data sources, including Wikipedia, arXiv, Github, Books, StackExchange, C4, and CommonCrawl^32^. The ViT was pre-trained on 2.3 billion images obtained from the web as part of the LAION-5B dataset^35^. The combined Flamingo model was pre-trained for its perceiver samplers and cross-attention blocks on 141 million interleaved image-text documents and 353 million images^31^.

### Testing Medical Image Interpretation

To test the medical reasoning of the models and their ability to stratify medical images for downstream tasks, we use a method similar to recently published approaches ^36–39^, i.e., we present the respective images to the model along with a general prompt, e.g., “What can you see on this radiological image?”. We then extract the representation of the images in the model’s internal latent space and test whether these representations can be used for classification by a simple linear logistic regression model, see **Figure 2c**. This concept is called “probing the model” and tests whether the internal representation of the images is linearly separable, i.e. whether the LLM has allocated healthy and pathological images to separate regions of its high-dimensional space.

### CLIP as a Comparison Model

We used OpenAI’s CLIP (Contrastive Language-Image Pre-training) as a benchmark to evaluate Flamingo’s performance. CLIP, specifically the CLIP-ViT-B/32 model, is trained on a corpus of over 400 million Internet-sourced image-text pairs, providing robust “zero-shot” learning capabilities^40^. We use this baseline model in all tasks T1-T8. As a second baseline model, focused only on the pathology tasks, we employ PLIP (Pathology Language-Image Pre-training), which has been trained with contrastive learning specifically on pathology images sourced from X (formerly Twitter) and has recently been presented as a foundational model with state-of-the-art performance in histopathology^11^.

### Image Pre-processing

Images larger than 1024×1024 pixels were downsampled to 1024×1024 pixels and underwent normalization relative to their maximum pixel value to ensure uniformity across the datasets. T2 and T8 required specific preprocessing: in T2, images of nuclei were processed according to the work of Huang et al^11^. The image was considered ‘malignant’ if the total number of neoplastic cells was more than ten and covered more than 30% of the total cells. Images were considered ‘benign’ if no neoplastic cells were present. This resulted in 2,866 malignant images and 3,368 benign images. For T8, knee radiographs were preprocessed to include only a 140 mm×140 mm region using a pre-trained hourglass network reported by Tiulpin et al^41^.

### Computational Resources

We use four NVIDIA A6000 (48GB) GPUs on a local server system to probe the models. To train the logistic regression model on the internal probes of Flamingo activations, an NVIDIA RTX 3090 (24GB) GPU was used.

### Evaluation and Statistical Analysis

For T3 to T8, the performance of the classifiers was evaluated by the area under the receiver-operator curve (AUC). For T1 and T2, the classification performance was evaluated by the F1 score according to Huang et al^11^. Standard deviations (SDs) and P values were calculated using bootstrapping with 1,000 replicates and paired 2-tailed t-tests.

## Data availability

The NEJM challenge questions are available to the public via: https://www.nejm.org/image-challenge. The validation datasets are publicly available and can be accessed from the following: Kather Colon (https://zenodo.org/record/1214456); PanNuke (https://warwick.ac.uk/fac/cross_fac/tia/data/pannuke); ISIC-2018 (https://challenge.isic-archive.com/data/#2018); ISIC-2020 (https://challenge.isic-archive.com/data/#2020); EyePACS Diabetic Retinopathy Detection (https://www.kaggle.com/c/diabetic-retinopathy-detection/); APTOS-2019(https://www.kaggle.com/c/aptos2019-blindness-detection); AIROGS (https://zenodo.org/records/5793241); ODIR-2019 (https://odir2019.grand-challenge.org/Download/); PadChest (https://bimcv.cipf.es/bimcv-projects/padchest/); OAI (https://nda.nih.gov/oai/query-download); MOST (https://most.ucsf.edu/multicenter-osteoarthritis-study-most-public-data-sharing).

## Code availability

The source codes can be accessed at https://github.com/peterhan91/Multimodal-Probes. The weights of open-sourced Flamingo models can be downloaded via https://huggingface.co/HuggingFaceM4/idefics-80b-instruct and https://huggingface.co/HuggingFaceM4/idefics-9b-instruct. OpenAI’s CLIP model can be downloaded via https://huggingface.co/openai/clip-vit-base-patch32. Inferencing of multimodal LLMs was performed using Huggingface transformers library (v.4.34.0.dev0, https://huggingface.co/docs/transformers/index) and PyTorch (v.2.0.1, https://pytorch.org/). Analysis of LLM’s representations was performed using Python (v.3.9.17, https://www.python.org/), scikit-learn (v.1.3.0, https://scikit-learn.org/stable/), and SciPy (v.1.11.1, https://scipy.org/).

## Results

### Accuracy in a Complex Diagnostic Challenge

First, we analyzed Flamingo-80B’s performance in answering clinical vignette questions from the NEJM challenge to mimic direct human-machine interaction through text. When testing on 929 cases, Flamingo-80B’s primary diagnosis matched the correct diagnosis in 40.4% (375 of 929) of cases (**Figure 1d**). When the model was prompted three times in succession, it included the correct diagnosis in 54.3% (504 of 929) of cases, as determined by stochastic top-K sampling with T=1.0 and top k=50. Notably, Flamingo-80B’s performance outperformed guesswork at various levels of difficulty, except for the most difficult category (**Figure 1d**). In **Figure 1a-c**, we illustrate selected Flamingo-80B responses and the explanation as provided by the model. These results highlight Flamingo-80B’s ability to provide medical insight and to integrate medical knowledge, albeit with the need for careful interpretation and validation in real-world settings.

### Systematic Testing in Histopathology, Dermatology, Ophthalmology, and Radiology

To put the vision-language models’ performance in a wide range of medical disciplines in context to existing pre-training methods, we presented image data with textual prompts to Flamingo-80B and Flamingo-9B, as well as using OpenAI’s CLIP as a benchmark.

### Vision-Language Models are Concept Encoders in Pathology

We investigated if histopathological images can be stratified by vision-language models and tested two tasks: classification of colorectal tissue and classification of nuclei. The colorectal tissue classification task (T1) focused on classifying tissue into nine categories based on hematoxylin & eosin-stained histologic images from a human colorectal cancer (CRC) cohort. In this task, a linear classifier was trained on internal activations obtained from multimodal LLMs and the CLIP model, analyzing a total of 7,158 histopathological image patches. The results showed that Flamingo-80B’s internal representations achieved a higher average F1 score of 0.892 as compared to the CLIP method, which scored 0.764. Notably, Flamingo-80B also outperformed the domain-specific foundation model developed by Huang et al.^11^, which was pre-trained on Twitter, with an F1 score of 0.892 versus 0.877. Detailed results for the different categories can be found in **Figure 3a-i**.

**Figure 3:**
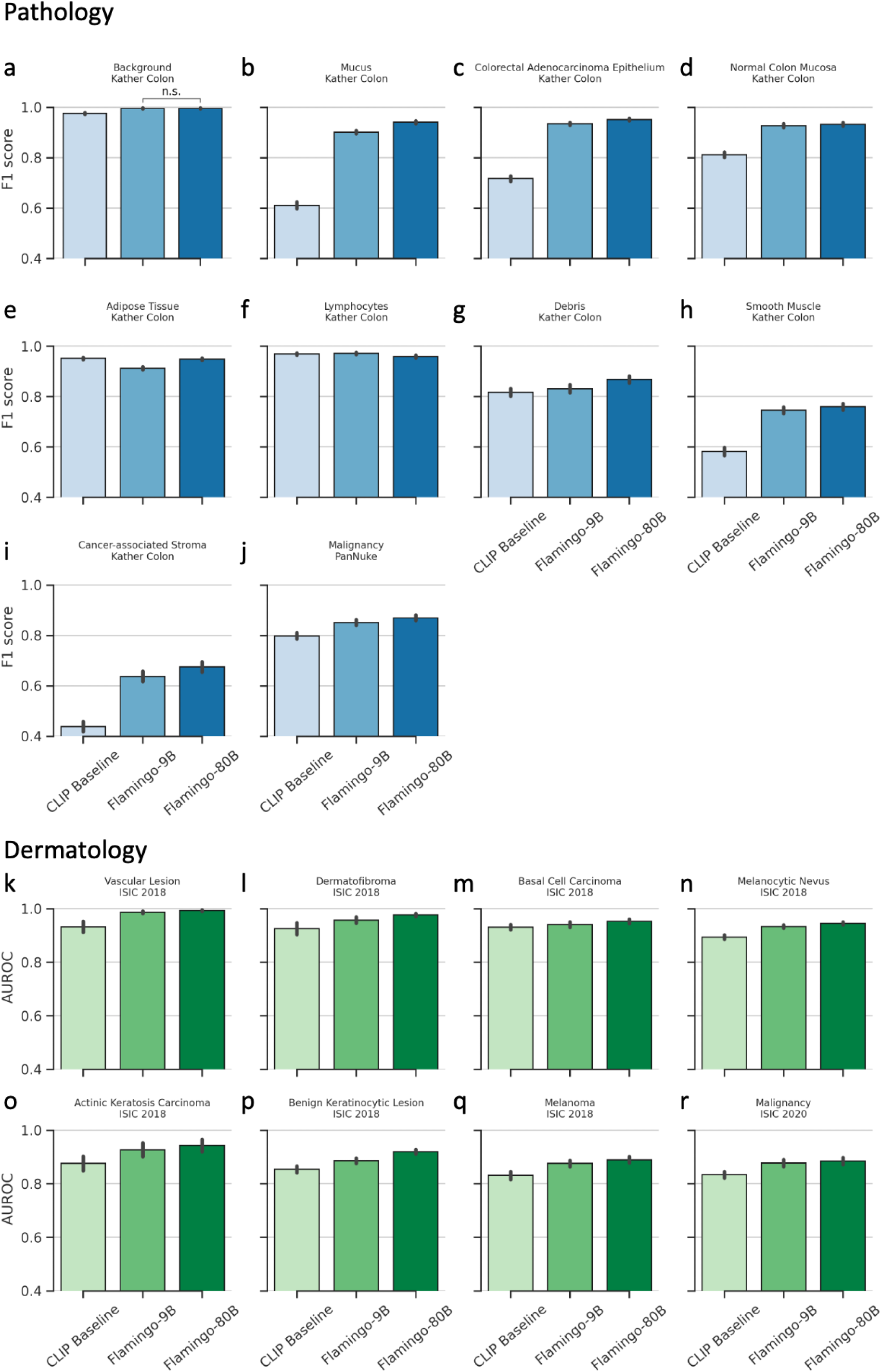
Performance in histopathological and dermatological image classification. (a-j) F1-score when classifying tissue type in task 1. Linear probes are fine-tuned on each dataset (Kather Colon and PanNuke) and evaluated on a hold-out test set. (a) to (i): classification of nine tissue types from colorectal cancer patients using image data from the Kather Colon dataset. (j): Malignancy classification in the PanNuke dataset in task 2. (k-r) AUC when classifying skin lesions. The probes are trained on the multimodal LLM’s internal representations to predict the type of skin lesions (k-q) and malignancy (r). The center of each bar represents the mean of the metrics (F1 and AUC) and the error bars indicate the SDs. SDs and P-values are calculated using bootstrapping and paired, two-tailed t-tests.

In the nuclear classification task (T2), our goal was to discriminate between benign and malignant cases among samples from 19 different organs using the PanNuke dataset (**Figure S6**). By applying a linear classifier to the internal activations derived from both multimodal LLMs and the CLIP model, Flamingo-80B demonstrated superior performance. Specifically, its internal representations yielded a consistently higher F1 score of 0.870 (95% CI: [0.847 to 0.891]) compared to the baseline CLIP method’s 0.797 (95% CI: [0.774 to 0.821]) (t-statistic=139.7, P<0.001), as detailed in **Figure 3j**. These results collectively confirm the advanced capabilities of multimodal LLMs over traditional pre-training methods in histopathology, even matching the accuracies of specialized foundation models that rely on domain-specific data.

### Vision-Language Models can Interpret Dermatological Images

We investigated if photos of skin lesions can be stratified by vision-language models and utilized two tasks to compare the performance of Flamingo-80B to CLIP pre-training: The first skin lesion task (T3) involves the multiclass classification of dermatological images into seven classes: melanoma, basal cell carcinoma, actinic keratosis carcinoma, melanocytic nevus, benign keratinocytic lesions, dermatofibroma, and vascular lesions. After training the linear classifier on the internal activations extracted from multimodal LLMs and the CLIP model Flamingo-80B’s internal representations resulted in a consistently higher AUC as compared with the baseline CLIP method in all seven classes, see **Figure 3k-q** for a more detailed breakdown (P<0.001 for all).

In the second skin lesion task (T4) on a separate dataset the models classified 33,126 dermatological images into malignant or benign lesions. Following the same architecture as above, Flamingo-80B achieved a significantly higher AUC on this task than CLIP (0.885, 95% CI: [0.859 to 0.909] vs. 0.834, 95% CI: [0.810 to 0.857], P<0.001), see **Figure 3r**. Together, these results show that vision-language models that have been pre-trained on non-domain-specific text- and image-data can differentiate between photos of dermatological lesions.

### Vision-Language Models are Generalist Image Interpreters in Ophthalmology

We performed additional experiments with two datasets of fundoscopic images to compare the vision-language models’ performance to pre-training with CLIP.

T5 focuses on the detection of diabetic retinopathy using over 90,000 fundus photographs in the US and India. Flamingo-80B shows superior performance in grading diabetic retinopathy (see **Figure 4**), especially in detecting proliferative and severe diabetic retinopathy (**Figure 4a, b**), achieving state-of-the-art results (AUC=0. 949, 95% CI: 0.939 to 0.958; and AUC=0.903, 95% CI: 0.889 to 0.917) and significantly outperformed the baseline CLIP model (AUC=0.883, 95% CI: 0.870 to 0.896 and AUC=0.826, 95% CI: 0.808 to 0.846; P<0.001 for both classes). Performance in detecting mild diabetic retinopathy is lower for all three models (Figure 4d), possibly due to class imbalance and labeling ambiguity, with Flamingo-80B performing best with an AUC of 0.629 (95% CI: 0.612 to 0.644).

**Figure 4:**
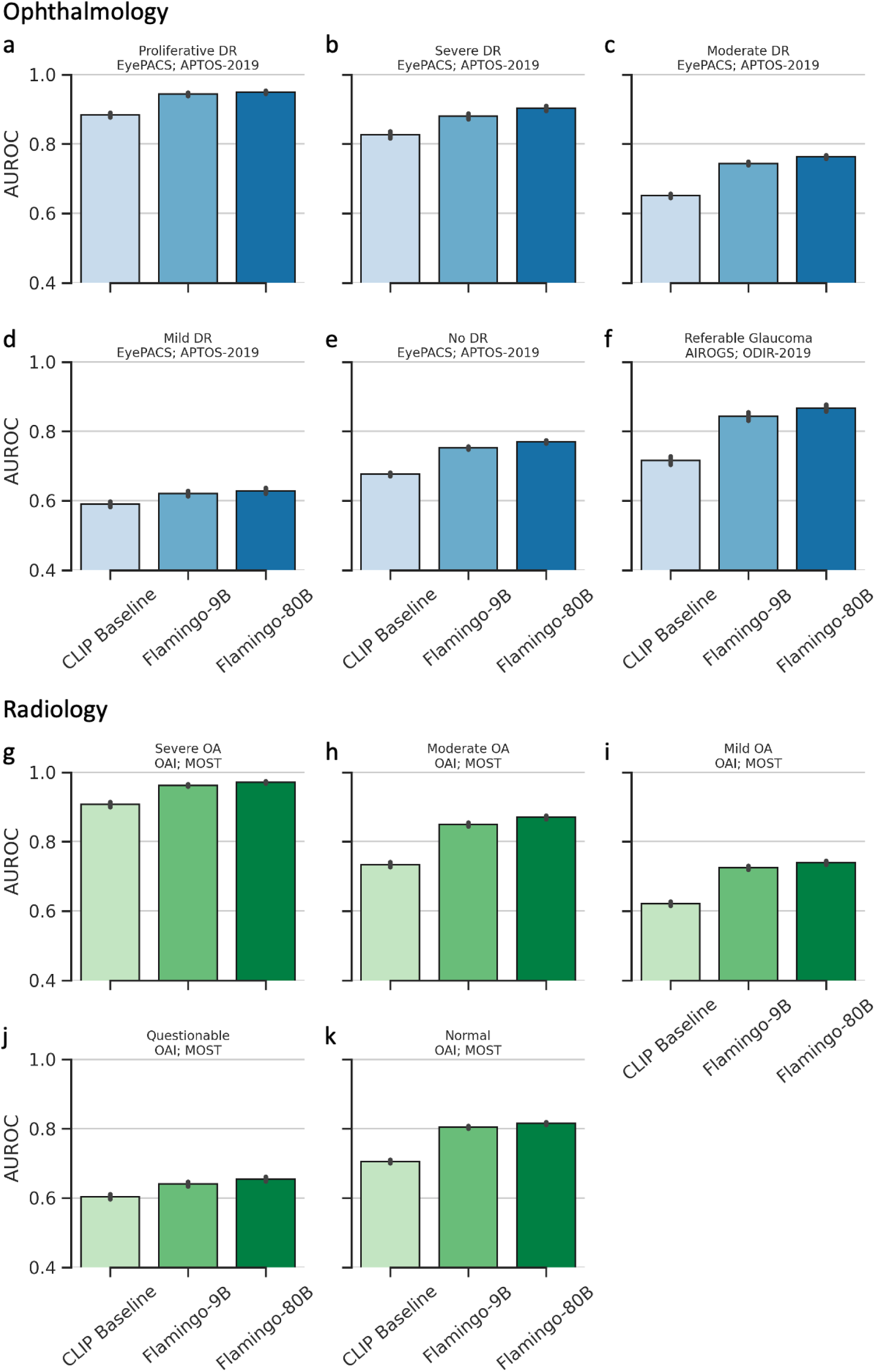
Performance in ophthalmological and radiological image classification. (a-e): Grading of diabetic retinopathy (DR). Linear probes are adapted to the EyePACS dataset by fine-tuning and evaluated on a hold-out test set to differentiate different stages of DR, such as proliferative DR, mild DR, and no DR eyes. (f): Classification of referrable glaucoma. (g-k) Performance in OA diagnosis based on knee radiographs. The center of each bar represents the mean AUC, and the error bars indicate the SDs. SDs and P-values are calculated using bootstrapping and paired, two-tailed t-tests.

T6 addresses another significant visual impairment cause, glaucoma, assessed in a large patient cohort from Beijing, China, comprising 3,500 individuals^42^. Here again, the probe trained on the Flamingo-80B activations showed superior performance in AUC (0.868) compared to both its smaller variant, Flamingo-9B (AUC: 0.843; P<0.001), and the baseline CLIP model (AUC: 0.716; P<0.00, **Figure 4f**).

Together these results show the general applicability of vision-language models to previously unseen images in ophthalmology underlining their potential for the development of generalist image interpreters.

### Vision-Language Models can find Radiological Abnormalities

We performed two experiments with radiological images to test the vision-language models’ identification of radiological findings and compare their performance CLIP.

The chest X-ray classification task (T7) aims at allocating 54 radiographic findings to chest X-rays from the PadChest dataset. We utilized 94,658 chest X-rays of which 27.9% were labeled manually by board-certified radiologists. A subset of 7,943 manually labeled chest X-rays was set aside for testing. After training the linear classifier on the internal activations of the multimodal LLMs, Flamingo-80B led to an AUC of at least 0.90 in 7 findings and of at least 0.70 in 40 findings. CLIP achieved these AUC thresholds in none and only 6 findings, respectively, see **Figure 5**.

**Figure 5:**
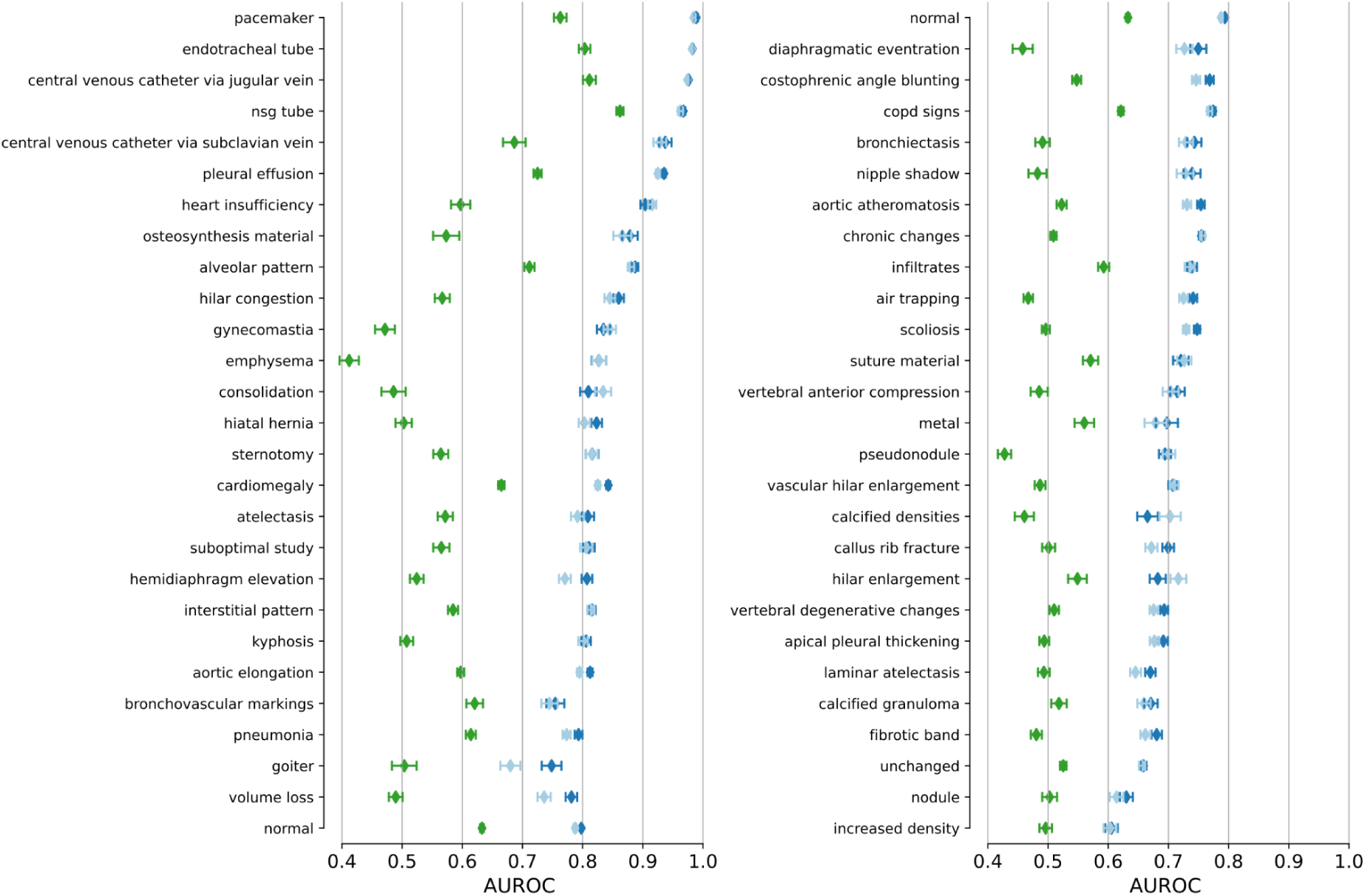
Detection of imaging and radiological findings on PadChest radiographs. Mean AUC and SD are shown for each finding with more than 50 entries in the PadChest testing cohort. The top 27 imaging findings are shown in the left panel and the remaining imaging findings are shown in the right panel. Flamingo-90B (green) consistently achieves higher AUC than CLIP (blue).

T8 investigates the performance of diagnosing osteoarthritis (OA) in knee X-rays. OA was graded based on manual labels by board-certified radiologists.^4^ Again training a linear model on the internal activations led to the superior performance of Flamingo-80B in severe OA (0.971, 95% CI: 0.965 to 0.976), moderate OA (0.870, 95% CI: 0.860 to 0.880), and no OA (0.815, 95% CI: 0.807 to 0.824). CLIP’s performance was consistently lower with an AUC of (0.907, 95% CI: 0.894 to 0.920) in severe OA, (0.734, 95% CI: 0.720 to 0.748) in moderate OA, and (0.706, 95% CI: 0.696 to 0.715) in no OA, see **Figure 4g-k**.

Together these results demonstrate that vision-language models can identify a wide range of radiological findings without having been specifically trained for this task.

### Vision-Language Models are data efficient

Our goal was to determine whether LLMs’ inherent knowledge and inference capabilities could facilitate the development of AI models using a reduced number of labels. To this end, we conducted a series of label efficiency experiments. These experiments were designed to determine the minimum amount of training data and labels required for LLMs to achieve specific performance benchmarks on various medical tasks.^10^

Our results were particularly striking with Flamingo-80B. Using only 10% of the training data, Flamingo-80B was able to retain good performance across four medical disciplines. Specifically, it maintained 95.8% (comparing an F1 score of 0.855 with 10% data to an F1 score of 0.892 with 100% data), 94.3% (comparing an AUC of 0.892 with 10% data to an AUC of 0.945 with 100% data), 95. 2% (comparing an AUC of 0.764 with 10% data to an AUC of 0.803 with 100% data) and 94.7% (comparing an AUC of 0.767 with 10% data to an AUC of 0.810 with 100% data) of its peak performance in pathology, dermatology, ophthalmology, and radiology, respectively. Detailed results of these findings are shown in **Figure 6**.

**Figure 6:**
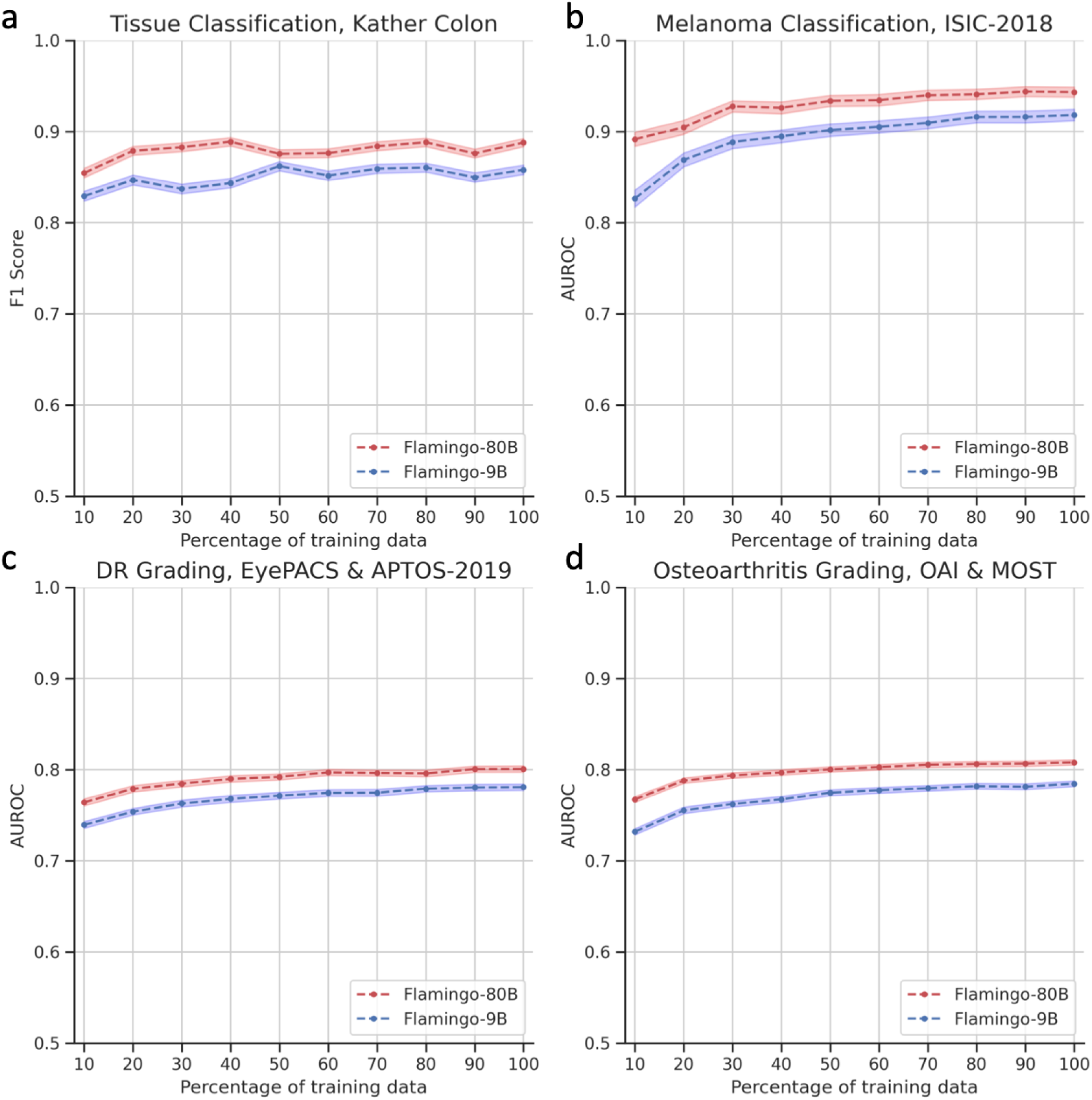
Robustness of our approach to data scarcity. In all four tasks, the performance of Flamingo-80B is robust to reducing the training data. Tuning on only 10% of the training data, we maintained 95.8%, 94.3%, 95.2%, and 94.7% of the best performances in the pathology, dermatology, ophthalmology, and radiology tasks, respectively. For tasks with multiple classification subtasks, we give the mean AUC. SDs is given as colored bands.

These results suggest that the knowledge and inference capabilities embedded in multimodal LLMs are highly effective, enabling the development of AI models with limited amounts of labeled data.^8^ This feature of LLMs holds great promise for applications where large labeled datasets are not readily available.

## Discussion

Deep Learning methods have been applied to a vast variety of medical problems in the past decade and their performance has increased to such an extent that they now equal or excel clinical experts in dedicated narrow tasks^1,43,44^. However, models that are good at solving one task, but fail to generalize to other tasks are of limited use in daily clinical routine.

This has ushered a push towards more general foundation models that are envisioned to carry out a diverse set of tasks using very little or no task-specific labeled data^8^. First steps in this direction have been taken by groups that trained domain-specific foundation models in ophthalmology^10^ and histopathology^11^. However, the training of such models still requires access to very large domain-specific datasets comprising millions of images. This limits the development of such models to a few groups. Furthermore, these models can not be regarded as true general foundation models as their applicability is limited to their specific domain. In this work, we present evidence that large vision-language models can serve as the sought general foundation models. We show how these vision-language models can be used as medical image interpreter base models that can be fine-tuned to specific tasks with a fraction of the data necessary to train conventional deep learning models. This opens up new possibilities in the application of AI to medical problems: there is a large number of medical tasks for which no AI models have been developed so far due to the scarcity of data. With vision-language models as general image interpreters, these tasks may now be tackled which can ultimately benefit clinical routine.

We performed our experiments with publicly available models that have been trained on publicly available data in a transparent manner. We thus set ourselves apart from research on the proprietary model GPT4-Vision^45,46^ which is touted as the state-of-the-art foundation model However, not much is known about the internal architecture, the model size, or the training data of GPT4-Vision. Not least due to its proprietary nature, the extent to which it can be used to drive progress in medicine is thus severely limited. Rather we focus on the open-source vision-language model Flamingo-80B. We show that Flamingo-80B inherently possesses medical knowledge and excels at classification tasks without specialized training. For this, we performed an extensive evaluation of eight datasets from four medical specialties comprising more than 450,000 medical images and demonstrated the wide applicability of our findings.

Our findings suggest a reevaluation of the current approach to AI in medicine, where specialist models are trained for new applications. Instead, generalist vision-language models offer a versatile, cost- and data-efficient alternative to the development of multiple specialized models. In addition, their inherent knowledge and ability to process information from other domains can facilitate the linking of different domains within the medical field and the incorporation of existing knowledge^47,48^.

Our work has limitations and leaves room for future research. Specifically, we performed a proof-of-concept and focused solely on imaging information. Therefore, we did not investigate the fusion of imaging information with more complex textual information, such as patient reports or patient history. Second, the model exhibited hallucinations when answering some of the clinical vignette questions for the NEJM challenge. We provided examples in **Figure S3** and **Figure S4**. There is thus an urgent need for the development of safeguards against these hallucinations.

Third, the NEJM challenge questions are not a factual representation of the clinical workflow, but rather a vignette of clinical cases used to evaluate the LLM’s clinical reasoning skills. Follow-up studies are necessary to establish the real clinical use of such models. Most importantly, we used LLaMA as the LLM backbone. While there are more powerful models like GPT4-Vision by OpenAI and Gemini Ultra by Google, we were unable to test these due to their closed nature, but we anticipate that they, along with future open-source LLMs, will result in even better performing vision-language models.

## Conclusions

Large vision-language models that have been trained on publicly available data can serve as general medical image interpreters. They are data-efficient and publicly available, rendering them ideal for the development of new AI models in medical areas where the lack of large labeled datasets has so far been prohibitive.

## Acknowledgments

None.

## Author contributions

T.H., L.C.A., K.K.B., J.N.K., and D.T. devised the study concept, and D.T. performed the reader tests. T.H. wrote the code and performed the performance studies. T.H. and D.T. did the statistical analysis. T.H., L.C.A., K.K.B., J.N.K., and D.T. wrote the first draft of the manuscript. All authors contributed to correcting the manuscript.

## Online Supplement

**Figure S1:**
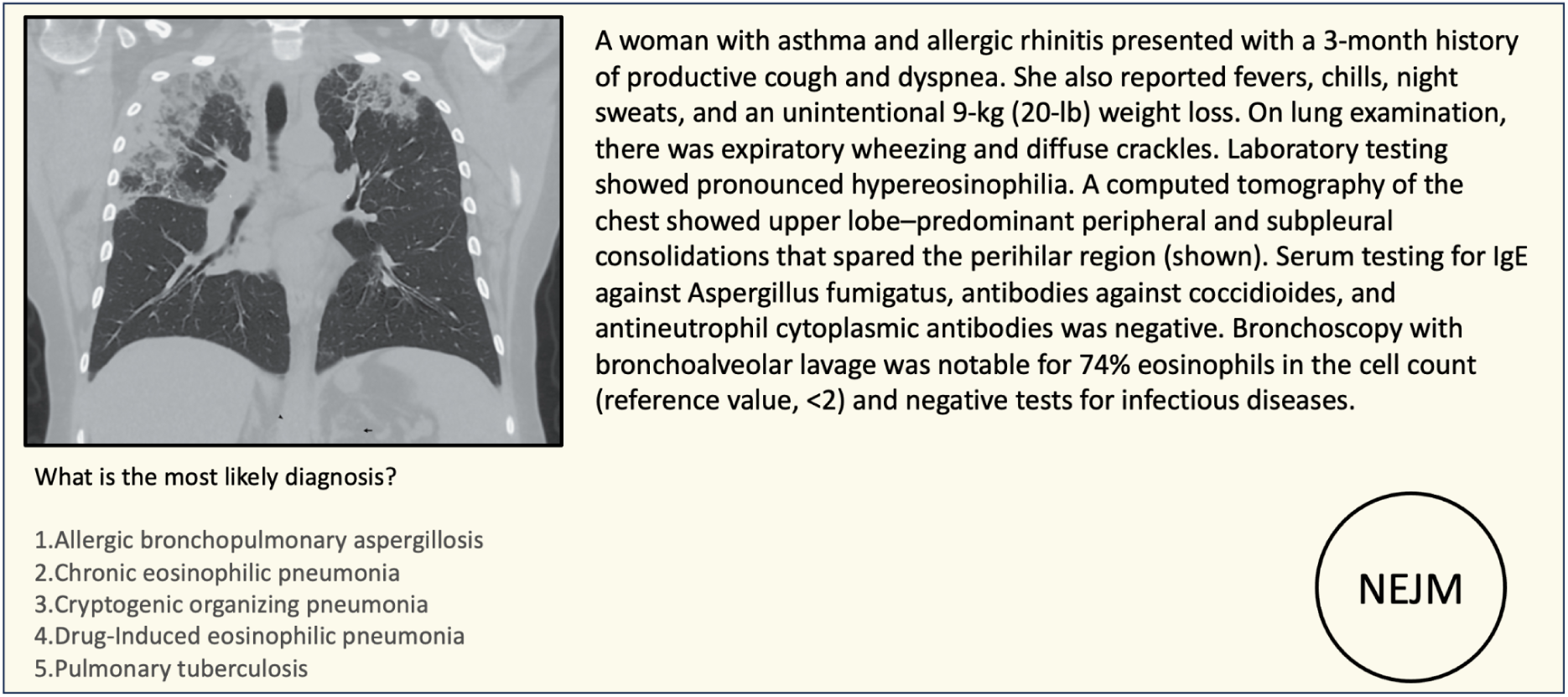
An illustrative example of the clinical case descriptions and answer choices from the “NEJM Image Challenge”.

**Figure S2:**
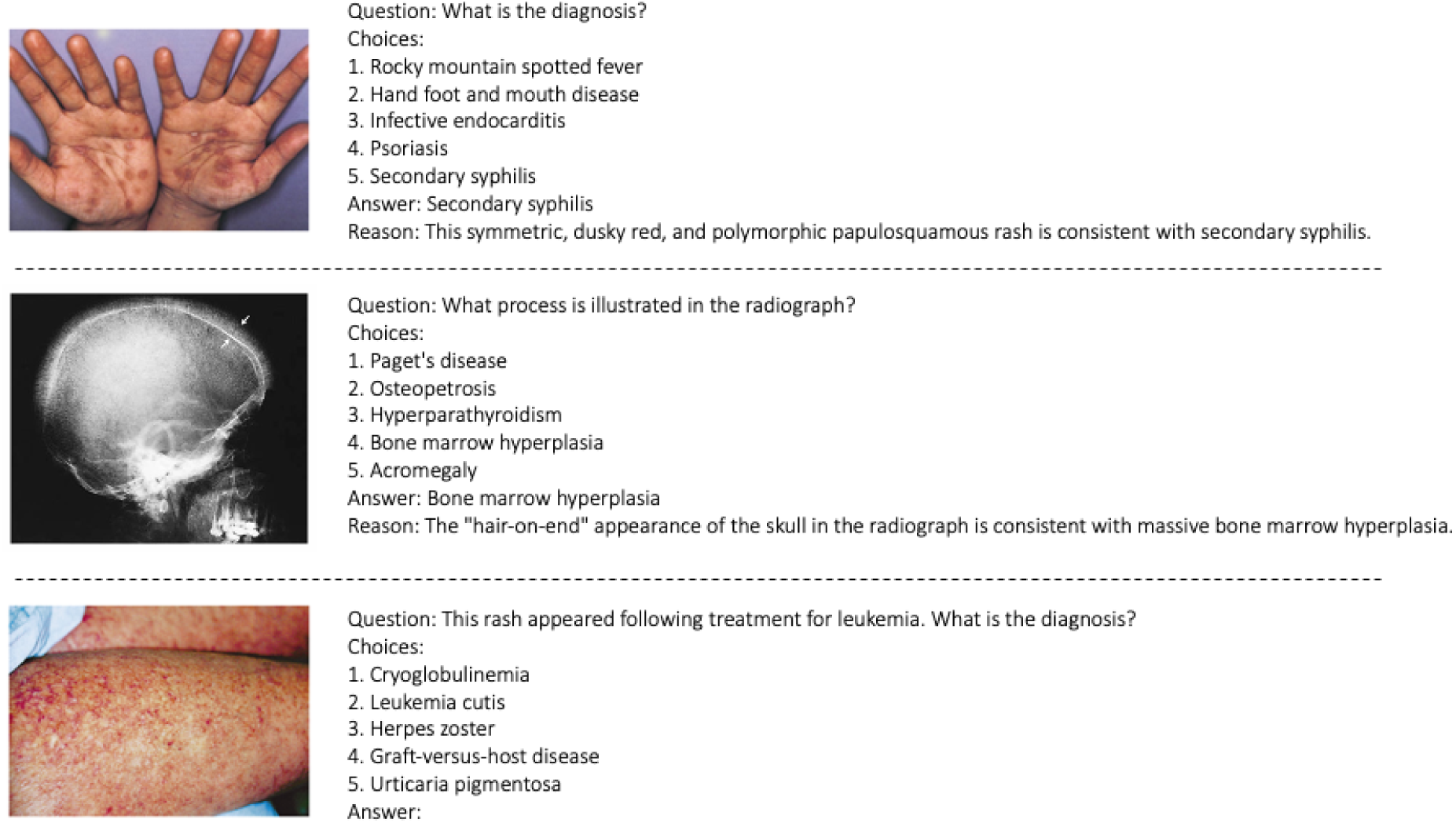
Two-shot example prompt used to query multimodal LLMs to answer NEJM Image Challenge questions.

**Figure S3:**
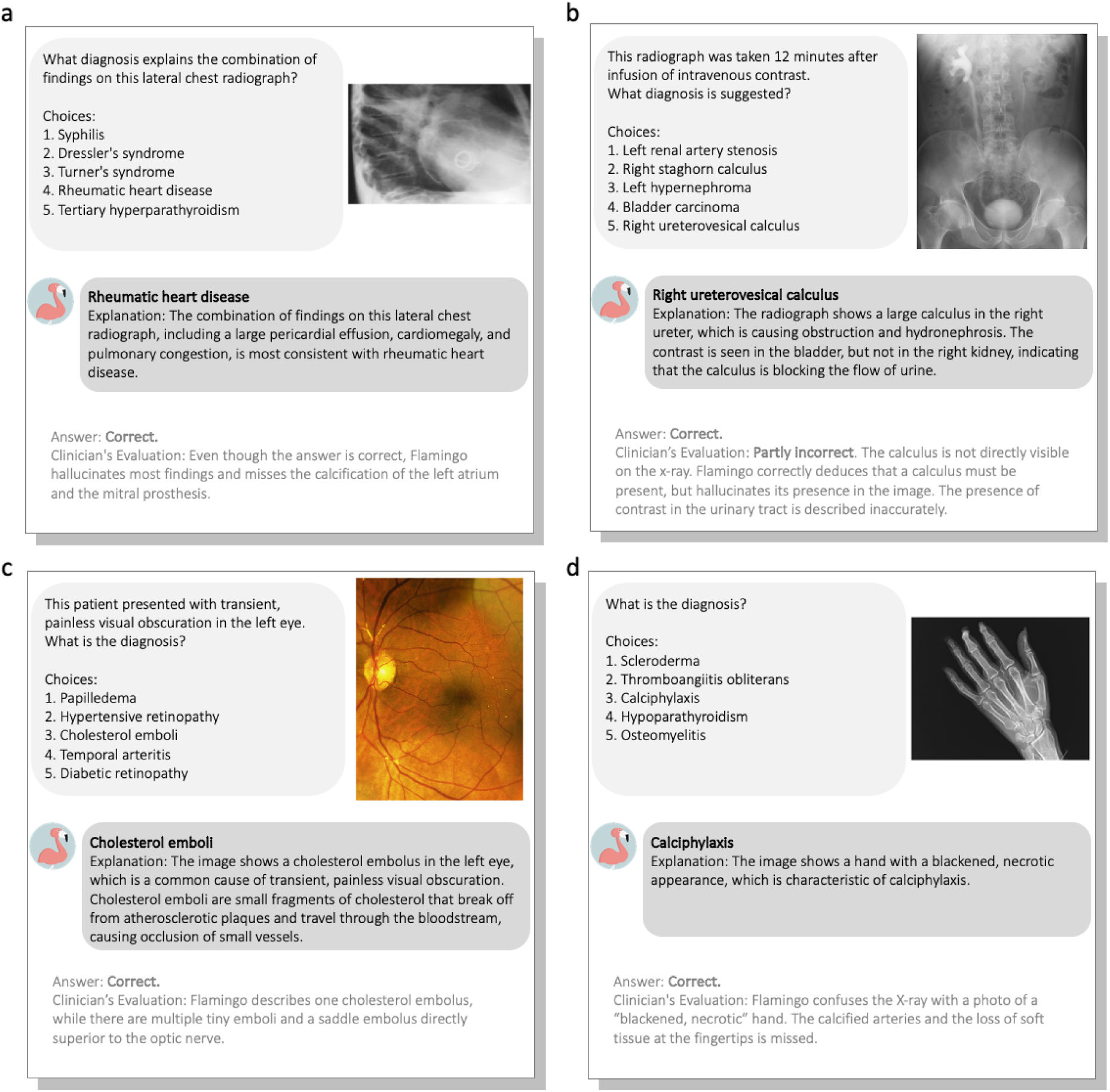
Cases from the NEJM Image Challenge with hallucinations. Flamingo-80B answered these questions correctly but reasoned incorrectly. We observed that multimodal LLMs can hallucinate strongly in certain medical cases such as (a), (b), and (c).

**Figure S4:**
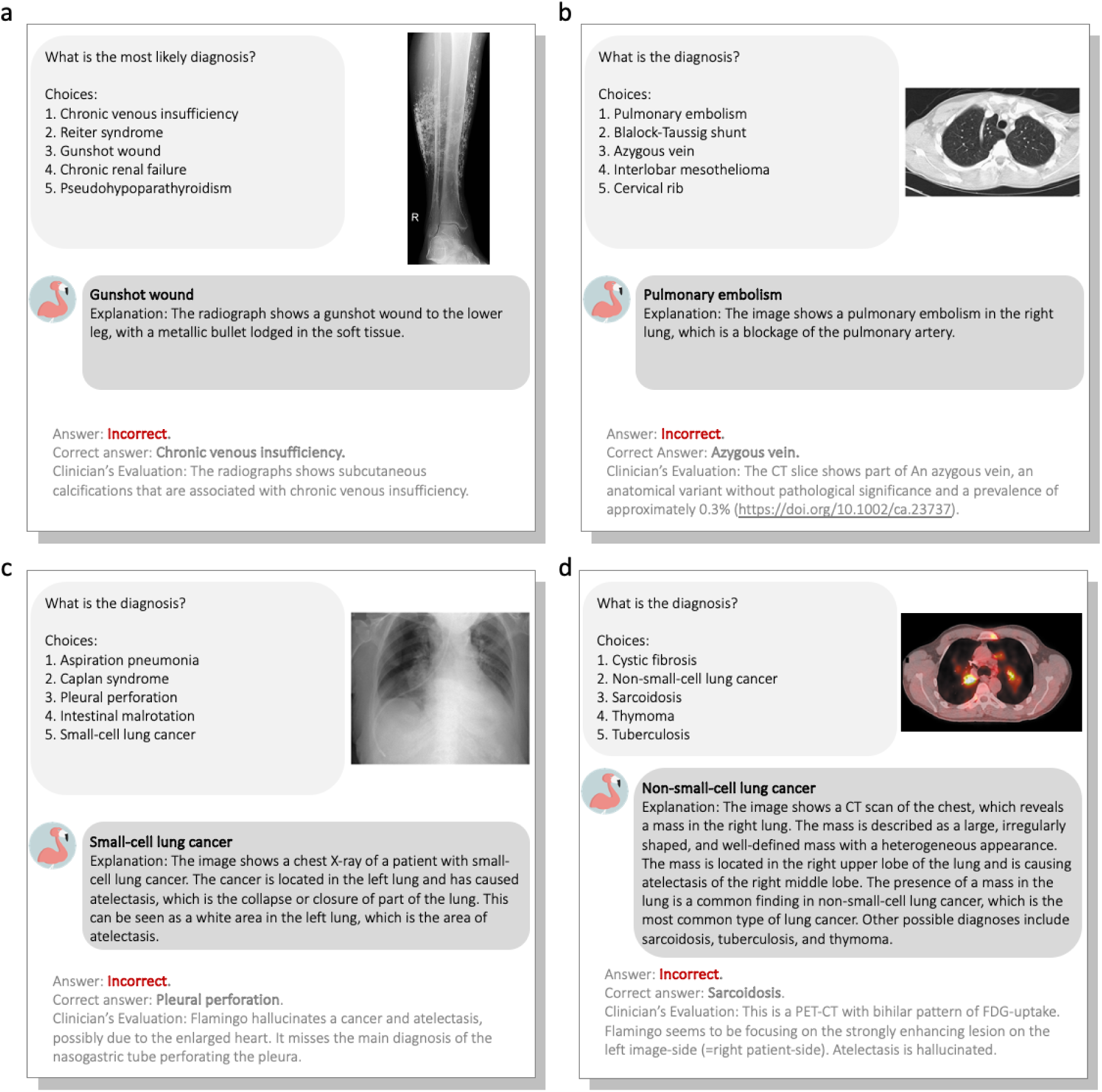
Selection of NEJM Image Challenge cases that were answered incorrectly. Flamingo-80B struggled to give the correct answer in these cases. We observe that Flamingo-80B mainly suffered from hallucinations (c) and (d) or misperceptions (a) and (b).

**Figure S5:**
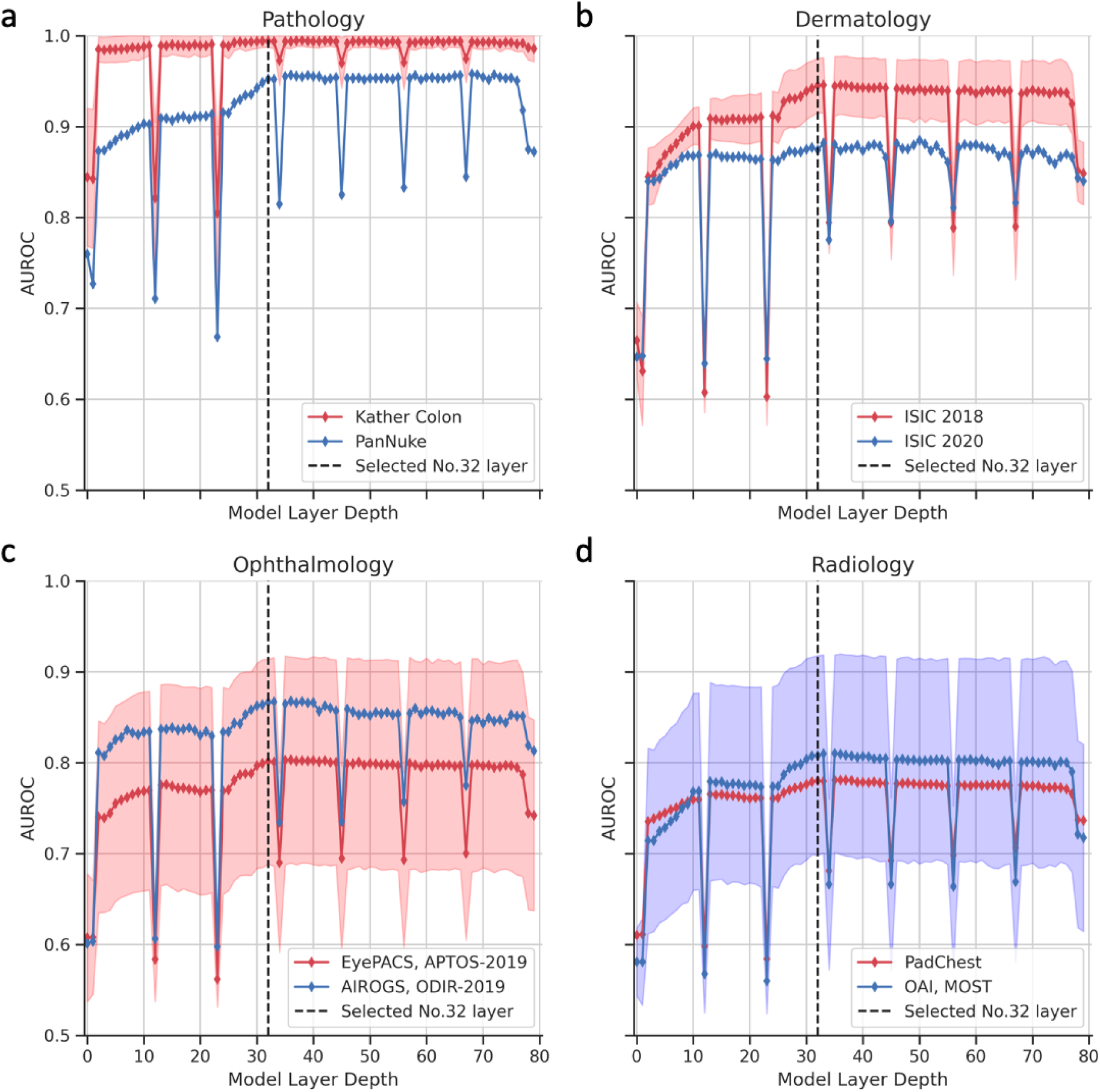
Testing the AUC for linear probes trained on each layer of the Flamingo-80B model. We select one layer (i.e., 32, highlighted in black, dashed lines) in a pre-experiment and then use it consistently for all subsequent experiments. In contrast to previously reported results,45 representations from the 80B multimodal LLM regularly fluctuate in quality across layers. We found that this phenomenon generalizes across evaluations in pathology (a), dermatology (b), ophthalmology (c), and radiology (d). The SDs of the AUCs are plotted in color bands, and the midpoints of the bands indicate the mean value of the AUC.

**Figure S6:**
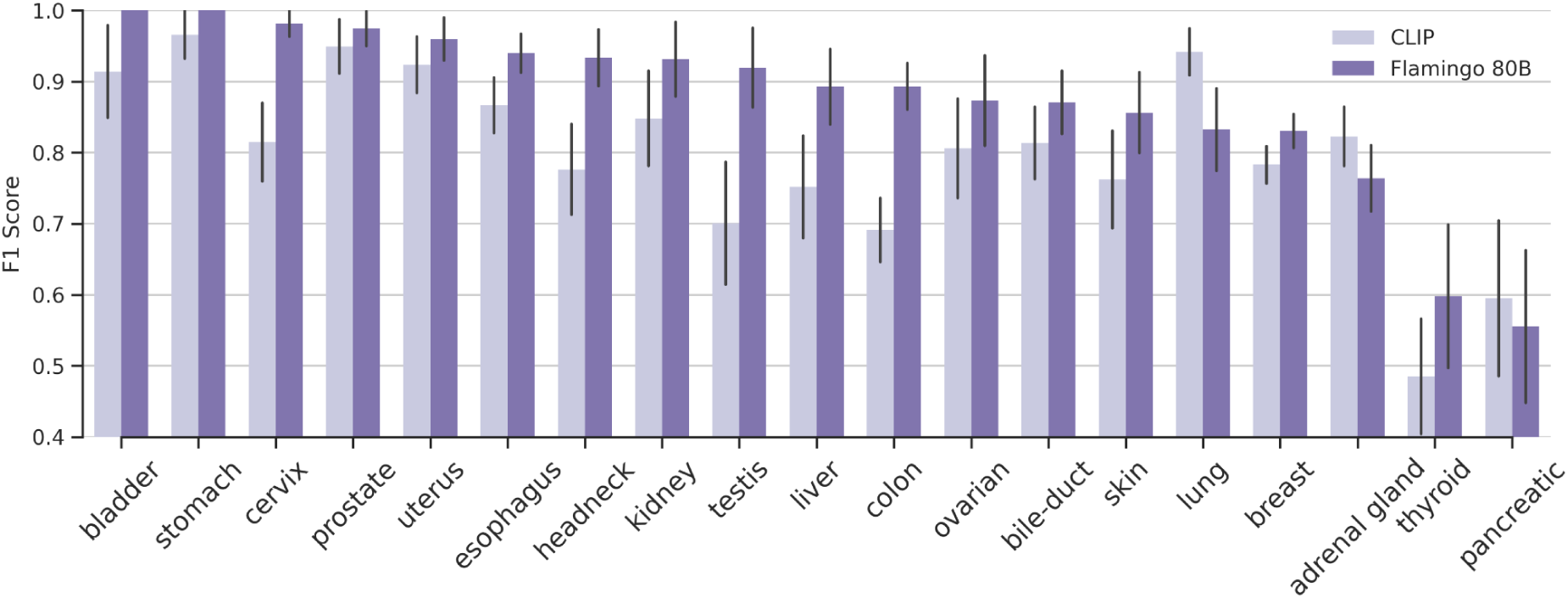
Evaluation of activation probes in the PanNuke dataset within each organ type.

